# Comprehensive Physical Exam versus Lung Ultrasound for Dyspneic Patients in the Emergency Department

**DOI:** 10.1101/2022.10.08.22280828

**Authors:** Michael Secko, Yuwen Cheng, Sonika Raj, Eshani Goradia, Lindsay Reardon, Henry C. Thode, Adam J. Singer

**Affiliations:** Department of Emergency Medicine, Renaissance School of Medicine at Stony Brook University, Stony Brook, New York, United States of America

## Abstract

**Objective:** Overreliance on technology has led to dwindling physical exam (PE) skills. We compared the diagnostic accuracy of a structured lung physical examination (L-PE) to structured lung ultrasound (LUS) in ED patients with undifferentiated dyspnea. We also examined the change in differential diagnosis and degree of certainty based on order and type of exam

**Methods:** This was a prospective, randomized, crossover study of a convenience sample of adult ED patients with undifferentiated dyspnea. Comprehensive L-PE and LUS were performed in random order followed by the other exam. An adjudication committee determined the final diagnosis based on all available data and served as the criterion standard. Primary outcome was diagnostic accuracy. A sample of 86 patients had 80% power to detect a 25% difference in diagnostic accuracy.

**Results:** A total of 102 patients were enrolled. Similar accuracies were found between L-PE and LUS for both COPD [75% (95% CI 65-83) vs. 76% (95% CI 67-84)] and asthma [87% (95% CI 79-93) vs. 87% (95 CI 79-93)]. LUS [81% (95 CI 72-88)] was slightly more accurate compared to L-PE [72% (95 CI 62-80)] for diagnosis of pneumonia but not statistically significant. For patients presenting with pulmonary edema, LUS was slightly [76% (95 CI 66-84)] more accurate than L-PE [73% (95 CI 63-81)], but not statistically significant. Finally, for detecting pleural effusions, L-PE [96% (95 CI 90-99)] was more accurate than LUS [82% (95 CI 73-89)].

**Conclusions:** The diagnostic accuracies of comprehensive lung physical examination and focused lung ultrasound were generally similar in ED patients with dyspnea and should be used concurrently to maximize diagnostic accuracy.

## INTRODUCTION

Since 1816, the stethoscope has been the symbol of a physician. For nearly 200 years, auscultation and physical examination have been the cornerstones of chest diagnostics.^1,2^ Countless educational texts have been published over centuries to guide young physician-trainees towards mastering the art of the physical exam.^3^ Now, modern diagnostic tools including point-of-care ultrasound are challenging this age-old paradigm.

The undifferentiated dyspneic patient is one of the more challenging cases for Emergency Physicians (EP). Critically ill patients with dyspnea and hypoxia frequently present to the emergency department (ED) and EPs must make rapid diagnostic decisions with limited clinical information. It is imperative to maximize benefit while avoiding unnecessary and potentially harmful testing and treatment strategies.^4,5^ The traditional clinical evaluation of a dyspneic patient typically entailed a history and physical exam (H&P) followed by routine chest radiography (CXR).^6^ Relying on the H&P has never been ideal, given that it is non-specific and often inconclusive, particularly for patient with chronic dyspnea.^7^ CXR findings are also often delayed, frequently misleading, and have a low sensitivity for pathology.^4-6,8^

Point-of-care ultrasound (POCUS), with its increasing portability and excellent imaging quality, is a rapid, reliable, and noninvasive tool for the diagnosis of dyspneic patients.^9^ Numerous studies have compared the performance of different diagnostic tools and have confirmed that POCUS is more accurate than both physical examination and CXR to diagnose patients presenting to the ED with undifferentiated dyspnea. ^6,10,11^ One of the arguments against the reproducibility of POCUS is that most studies are done by clinicians with above-average experience with sonography.^5^ Studies have shown, however, that as few as 15 quality cardiac and thoracic scans are enough to train clinicians and improve their confidence in their leading diagnosis. ^12,13^

Medical educators are now emphasizing US in the curricula for future health care providers and there is momentum towards replacing the stethoscope.^14,15^ The increasing availability of advanced technologies and sensitive diagnostic testing has contributed to a decline in physical exam skills of medical students, interns, and residents. ^16,17^ The recognition that physicians are relying more on technology and less on physical exam skills has been circulating since 1988.^2,18^ The few studies that have specifically compared PE versus US for respiratory conditions (e.g. dyspnea), however, are limited to auscultation with a stethoscope.^6,19,20^ Auscultation is only one of the many maneuvers that may be performed as part of a comprehensive PE. Other techniques include inspection, palpation, and percussion, which are rarely used by many physicians after medical school.

We compared structured comprehensive lung PE (L-PE) to structured lung ultrasound (LUS) as a diagnostic tool for patients presenting to the ED with undifferentiated dyspnea. To the best of our knowledge, there are no publications that explicitly compare a comprehensive L-PE with a LUS protocol. We further evaluated the change in differential diagnosis and degree of certainty as secondary end points, as well as patients’ satisfaction and confidence level with their ED assessment.

## METHODS

### 2.1 Study Design

This is a prospective, crossover superiority study comparing a structured PE to a structured US exam in convenient sample of patients who present to the ED with acute dyspnea. This study was reviewed and granted approval by our institutional review board. All patients gave written informed consent.

### 2.2 Study Setting and Population

The study took place between July 2018 to July 2019 in the emergency department of a suburban, university-based hospital with approximately 74,000 adult ED visits per year. The hospital has an ACGME accredited PGY1-PGY3 emergency medicine residency program. All study members were resident (PGY1 – 3) or attending level emergency physicians. To participate in the study, the resident physician had to have completed a 2-week ultrasound rotation which included a 60-minute didactic on lung ultrasound as well as performed and completed 25 quality assured lung ultrasounds. PGY-1 and PGY-2 were classified as junior level of training and PGY3 and attending were classified as senior level of training. Attending physicians had to be credentialed in point-of-care ultrasound, which requires completion of at least 25 quality assured scans meeting American College Emergency Physician’s ultrasound requirements for POCUS and signed off by Chairman of the department.

Patients were enrolled when one of the study investigators was present. Criteria for enrollment included hemodynamically stable adult patients (>18 years) with chief complaint (CC) of shortness of breath (SOB) or dyspnea with the capacity to give informed consent. Hypoxic (<90% Oxygen saturation) patients were enrolled if they were determined clinically stable and able to tolerate the exam. A legally authorized representative (LAR) was eligible to give consent for patients who lacked capacity. The study excluded patients under the age of 18, those requiring life-saving procedures, and patients who lacked the capacity to give informed consent and had no LAR present.

### 2.3 Study Protocol

The allocation of patients in our study are found in **Figure 1**. Study investigators screened electronic medical records (EMR) and identified potential subjects with a chief complaint of dyspnea. The treating or research physician performed an initial screening examination and stabilized patients if needed. The study investigators then obtained informed consent. After consent, patients were randomized to either structured L-PE followed by structured LUS (L-PE/LUS) or structured LUS followed by structured L-PE (LUS/L-PE) group. A Study Coordinator directed patients to complete the Baseline Demographic/Clinical Characteristic Form and Patient Survey.

**Figure 1.**
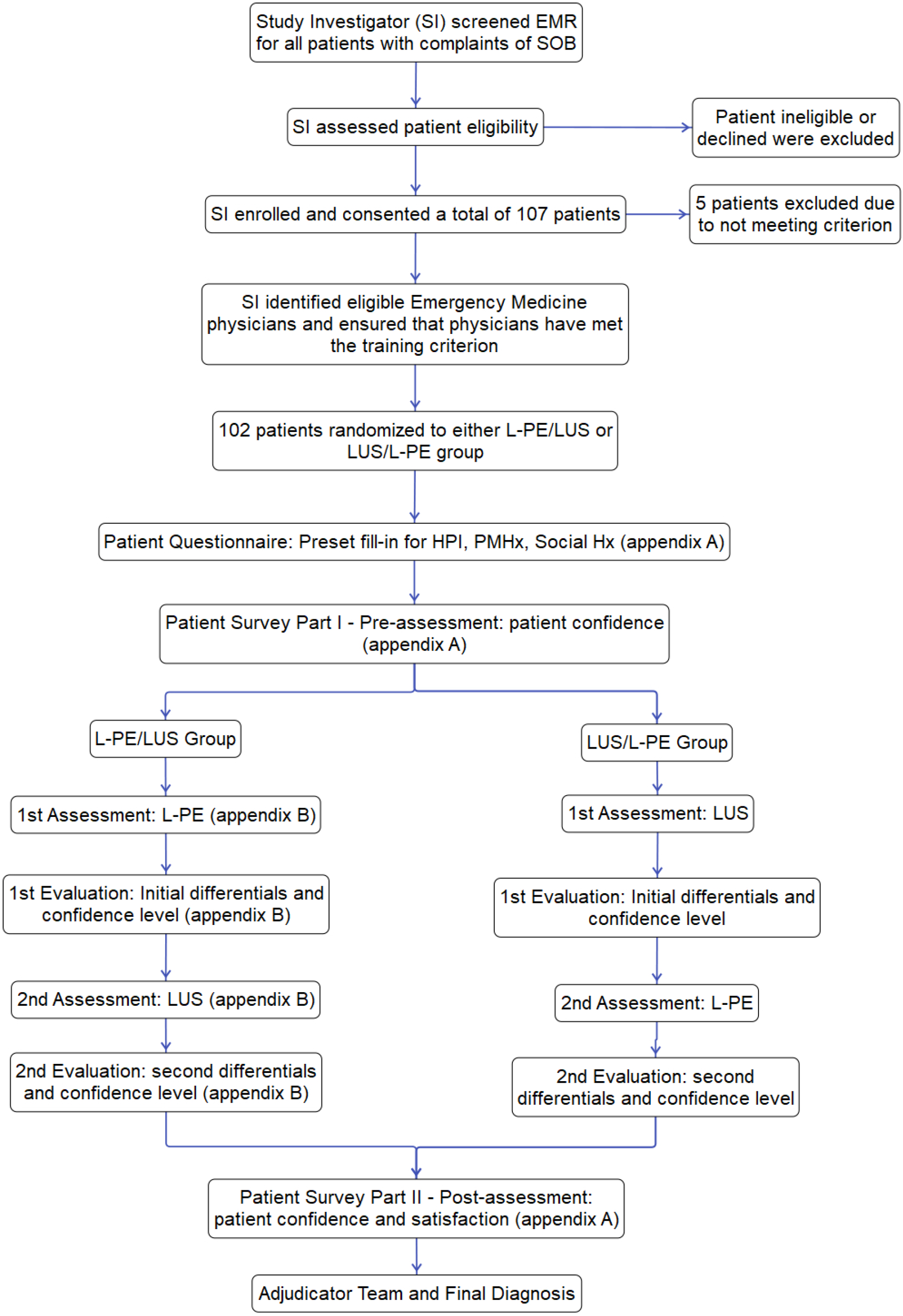
Flow diagram for allocation of patients

Study physicians were not blinded to the patient’s clinical history and were provided with patient’s Baseline Demographic/Clinical Characteristic Form and performed both the L-PE and LUS before any further ancillary or radiologic testing were performed or resulted. Physicians performed the first assigned assessment (L-PE or LUS) and completed the Structured Exam Form section pertaining to the conducted exam, which included clinical figures and differential diagnoses with degree of certainty. Afterward, physicians performed the second assigned assessment (LUS or L-PE) and completed the corresponding section in the Structured Exam Form **[Appendix]**. After both assessments, the study investigators directed patients to complete the Patient Survey again.

The investigated and measured diagnoses focused primarily on lung pathology, including asthma or chronic obstructive pulmonary disease (COPD), consolidation, pneumothorax, pleural effusion, and pulmonary edema. A write-in for other diagnoses not listed was available such as anemia, pericardial effusion, pulmonary embolism but was not the focus of this study since we were not examining or performing ultrasound of the heart.

All LUS were performed using a 5-1 MHz curvilinear array transducer. Sonographers used either a Mindray M9 (Mindray, Mahwah, NJ), Mindray TE7 (Mindray, Mahwah, NJ), or SonoSite Edge II (Fujifilm, Bothell, WA) ultrasound system to perform and record their images. All LUS exams were archived on the secure middleware software *QpathE* (British Columbia, Canada) and reviewed for quality assurance by an ultrasound fellowship-trained physician blinded to the clinical parameters.

An adjudication committee independently collected and reviewed each chart after the hospital course was completed. The committee was made up of 2 attending ultrasound trained EPs that were blinded to study results. Disagreements were managed by a third senior EP. They had access to the entire electronic record including all lab, echocardiography, radiologic results, admission notes, and hospital discharge summaries. Based on all available clinical data, the committee determined the final diagnosis, which served as the criterion standard. **[Appendix]**

### 2.4 Structured Lung Physical Exam

The study team created the structured lung and thoracic exam from Bates Guide to Physical Examination and History Taking, 12e. ^21^ The participating physicians were given a 10-minute didactic to refresh physical examination skills acquired during their medical school training. The exam consisted of 12 fields. The thorax was divided into 2 sections (anatomical right and left lung). Each hemithorax was separated into 3 zones. The anterior chest wall or zone 1 was delineated from the sternal border to anterior axillary line; the lateral chest wall or zone 2 bordered between the anterior and posterior axillary line; and the posterior chest wall or zone 3 consisted of the area between the posterior axillary line and mid-scapular line. Each section was divided into 2 partitions (upper and lower quadrant) **[Figure 2]**. Symmetrical comparisons were separately noted. Structured comprehensive L-PE included inspection, palpation, percussion, and auscultation in regions mentioned above. Techniques and standard characteristics were as described.^21^

**Figure 2.**
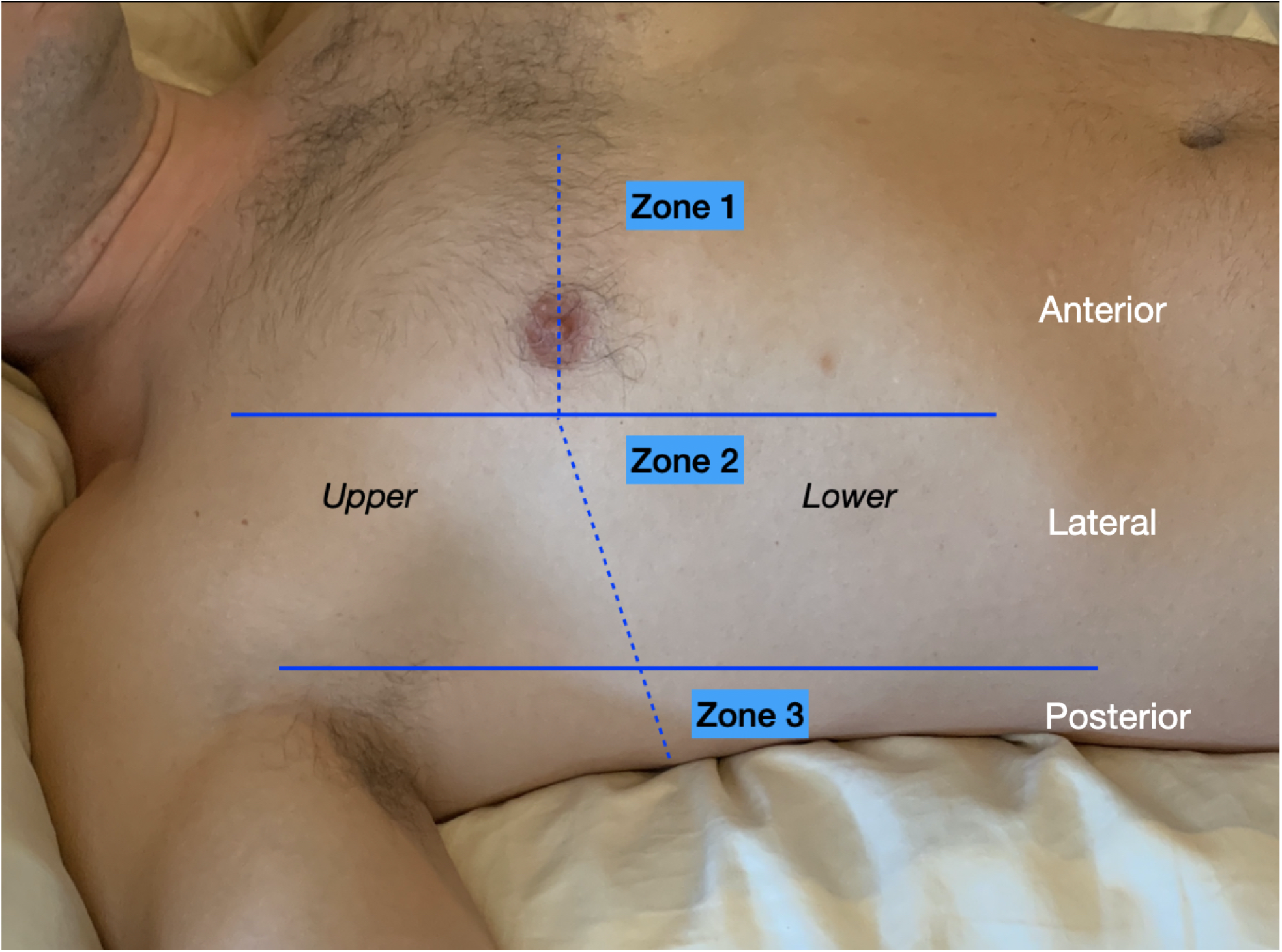
Structured Physical and Ultrasound Examination

### 2.5 Structured Lung Ultrasound Exam

We followed the structured lung ultrasound exam described in The BLUE Protocol.^22^ The exam consisted of 12 fields. The thorax was divided into anatomical right and left hemithorax. Each hemithorax was divided into the anterior, lateral, and posterior zones as described above. Each zone subdivided again into superior and inferior regions [Figure 2]. The lung ultrasound was classified into the following categories according to previously described criteria.^6,9,22,23^ Lung ultrasound artifacts include A-lines, which are horizontal lines repeating below the pleural line, and B-lines or comet tails, which are vertical lines extending from and perpendicular to the pleural line. Pathological B-lines obscure A-lines and reach the edge of the screen **[Fig. 3a]**.^24^ The absence of lung sliding or the presence of “lung point” defined a pneumothorax.^25^ Alveolar– interstitial syndrome was defined as the presence of equal to or greater than 3 B-lines in a given lung region **[Fig. 3b]**.^6^ Pleural effusion was defined as dependent, traditionally anechoic collection limited by the diaphragm and the pleura with respiratory movement of the lung within the effusion **[Fig. 3c]**.^6,26,27^ Consolidation was defined by 1) the presence of a “shred sign” or irregular border between consolidated and aerated lung, 2) hyperechoic punctiform images representative of dynamic air bronchograms, and/or 3) the tissue-like pattern called “hepatization” **[Fig. 3 d&e]**.^6,9,28^

**Figure 3a-e.**
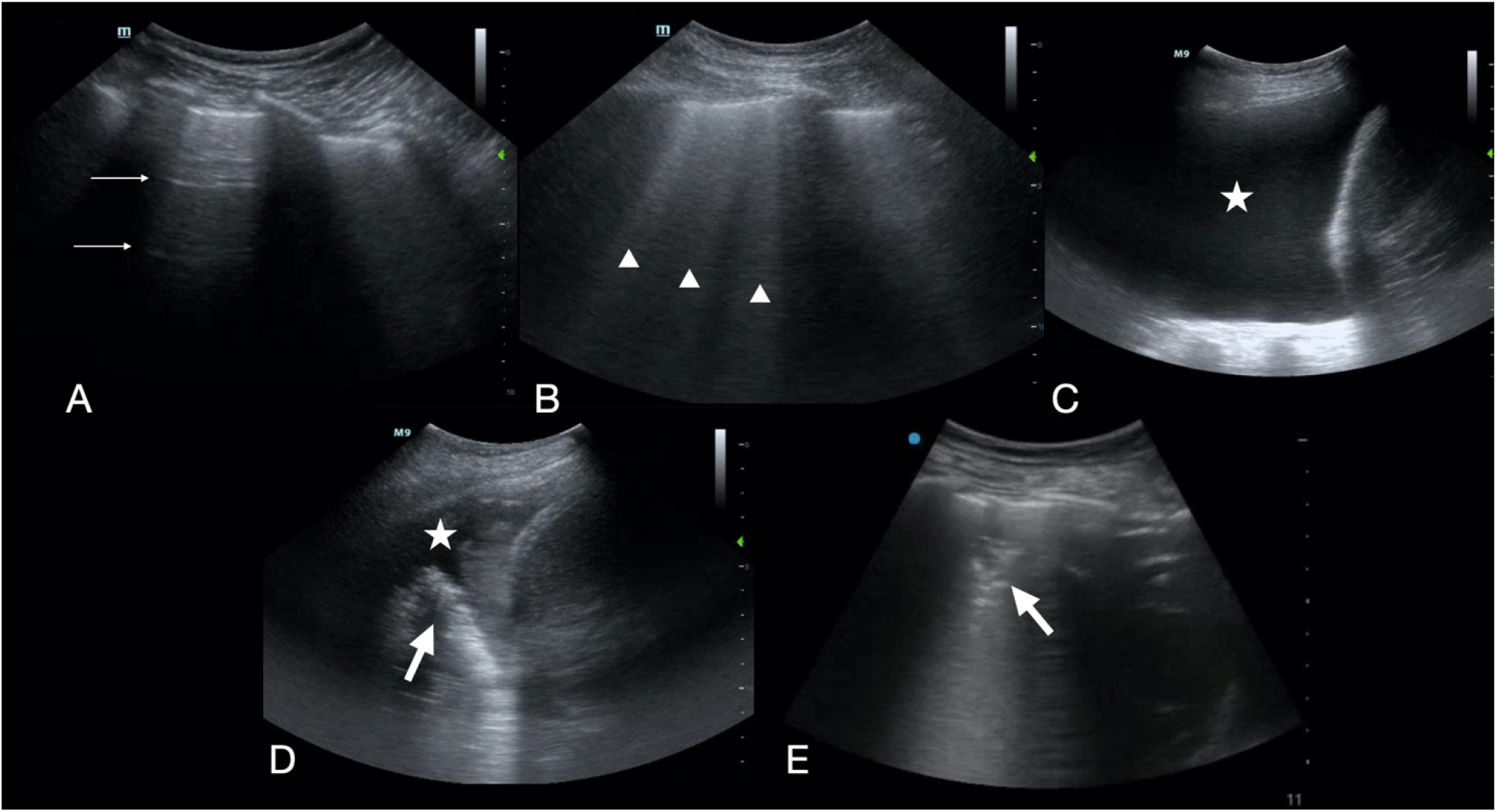
Lung ultrasound findings. (A) A-lines designated by arrows, (B) B-lines indicated by arrow heads, (C) pleural effusion designated by star, (D) pneumonia labeled by arrow with parapneumonic effusion designated by star, (E)

### 2.6 Outcomes

The primary outcomes were 1) the diagnostic accuracy of initial L-PE compared to the criterion standard in dyspneic patients presenting to the ED, 2) the diagnostic accuracy of the initial LUS compared to the criterion standard in dyspneic patients presenting to the ED, and 3) the diagnostic accuracy of L-PE compared to LUS. Secondary outcomes included 1) physician change in differential diagnosis between asthma/COPD versus pulmonary edema after the first versus second assessment, 2) physician change in degree of certainty in leading diagnosis after the first versus second assessment.

### 2.7 Statistical Analysis

Assuming the accuracy of structured thoracic PE is 65% ^8,29-31^ in diagnosing the cause of SOB (e.g., heart failure, COPD, pneumonia, pulmonary embolism), in order to achieve an 80% power to detect an absolute 25% increase in the diagnostic accuracy by ultrasound with a significance level <0.05, a sample of 43 patients was required in each group. Assuming a dropout rate of 10%, 102 patients were sampled in total. Statistical analyses were performed using SPSS version 26 (IBM, Armonk, NY). Patient baseline demographics were expressed as mean ± standard deviations. Diagnostic accuracy of initial structured L-PE or LUS compared to the criterion standard were represented as sensitivity, specificity, and diagnostic accuracy. Diagnostic accuracy between structured L-PE and structured LUS were compared using the chi-square test. Cohen’s kappa statistics were used to calculate the physician change in differential diagnosis after the first and second assessment, physician change in degree of certainty in leading diagnosis after the first and second assessment, and patient change in confidence level with L-PE and with LUS examination after the first and second assessment. Chi-Square testing was also used to analyze the comparison of patient satisfaction with medical care in general after intervention between L-PE/LUS and LUS/L-PE groups, the comparison of patient satisfaction with physical examination alone between L-PE/LUS and LUS/L-PE groups, and the comparison of patient satisfaction with ultrasound examination alone between L-PE/LUS and LUS/L-PE groups.

## RESULTS

### 3.1 Study Population

A total of 102 patients were enrolled. Patient demographics can be found in ***Table 1***. Fifty-seven patients (56%) were male and the mean age ± SD was 63 ± 18.5 years. Hypertension was the most common co-morbidity with 53 (52%) patients having the disease and 66 (65%) of the patients reported a history of smoking. Of those 102 patients, 50 patients (49%) had L-PE before LUS, and 52 patients (51%) had LUS before L-PE. A total of 23 EPs, 19 residents and 4 attending physicians participated in the study.

**Table 1.** Patient Demographics PE: physical examination, US: ultrasound, CHF: congestive heart failure, CAD: coronary artery disease, COPD: chronic pulmonary disease, DVT: deep vein thrombosis.

|  | PE first (n=50) | US first (n=52) | P value |
| --- | --- | --- | --- |
| Gender, n (%) |  |  | .41 |
| Males | 30 (60%) | 27 (52%) |  |
| Females | 20 (40%) | 25 (48%) |  |
| Mean age (SD) | 64 (18) | 62 (19) |  |
| History, n (%) |  |  |  |
| Diabetes | 12 (24%) | 10 (19%) | .63 |
| Hypertension | 26 (52%) | 27 (51%) | 1.00 |
| Renal disease | 3 (6%) | 7 (13%) | .32 |
| CHF | 11 (22%) | 14 (27%) | .65 |
| CAD | 10 (20%) | 12 (23%) | .81 |
| Asthma | 8 (16%) | 9 (17%) | 1.00 |
| COPD/emphysema | 14 (28%) | 16 (31%) | .83 |
| DVT/Pulmonary embolism | 4 (8%) | 4 (8%) | 1.00 |
| Cancer | 12 (24%) | 12 (23%) | 1.00 |
| Smoking | 35 (70%) | 31 (60%) | .31 |

### 3.2 Primary Outcomes

The adjudicated final diagnoses are noted in ***Table 2***. Thirty-five patients (34%) had either COPD or asthma for their final diagnosis, followed by 32 patients (31%) with pulmonary edema, while 20 patients (20%) were diagnosed with pneumonia. Forty-nine patients (48%) had other diagnoses not listed. With regard to the total number of final diagnosis, 69 patients (68%) had only one final adjudicated diagnosis while 33 patients (32%) had 2 or more diagnoses.

**Table 2.** Adjudicated Final Diagnosis Sums to more than 100% because of multiple diagnoses. COPD: Chronic obstructive pulmonary disease.

| *Adjudicated Final Diagnosis* | *Number* |
| --- | --- |
| COPD | 29 |
| Asthma | 6 |
| Consolidation | 20 |
| Pneumothorax | 0 |
| Pleural effusion | 8 |
| Pulmonary edema | 32 |
| Other | 49 |
| *Number of diagnoses* |  |
| 1 | 69 |
| 2 | 25 |
| 3 | 7 |
| 4 | 1 |

The overall accuracies of both physical exam and ultrasound for each diagnosis studied can be found in ***Table 3***. Similar accuracies were found between L-PE [75% (95% CI 65-83)] and LUS [76% (95% CI 67-84)] for COPD as well as asthma with [87% (95% CI 79-93) vs. 87% (95% CI 79-93)] for both L-PE and LUS. For diagnosing pneumonia, LUS [81% (95% CI 72-88)] was slightly more accurate compared to L-PE [72% (95% CI 62-80)], but not statistically significant. For detecting pleural effusions, L-PE [96% (95% CI 90-99)] was more accurate than LUS [82% (95% CI 73-89)]. With regard to pulmonary edema, LUS was slightly [76% (95% CI 66-84)] more accurate than L-PE [73% (95% CI 63-81)] but was not statistically significant.

**Table 3.**
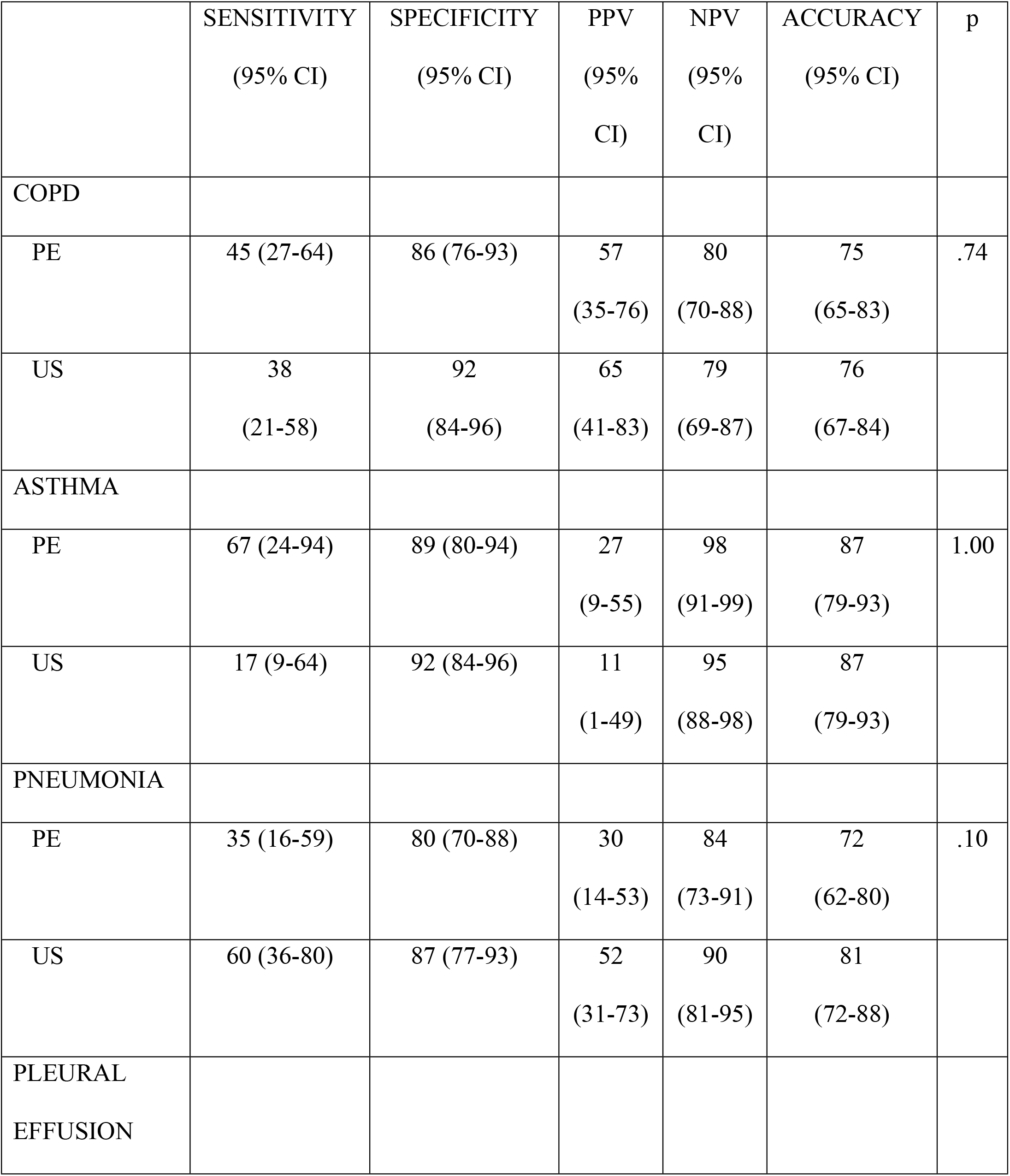
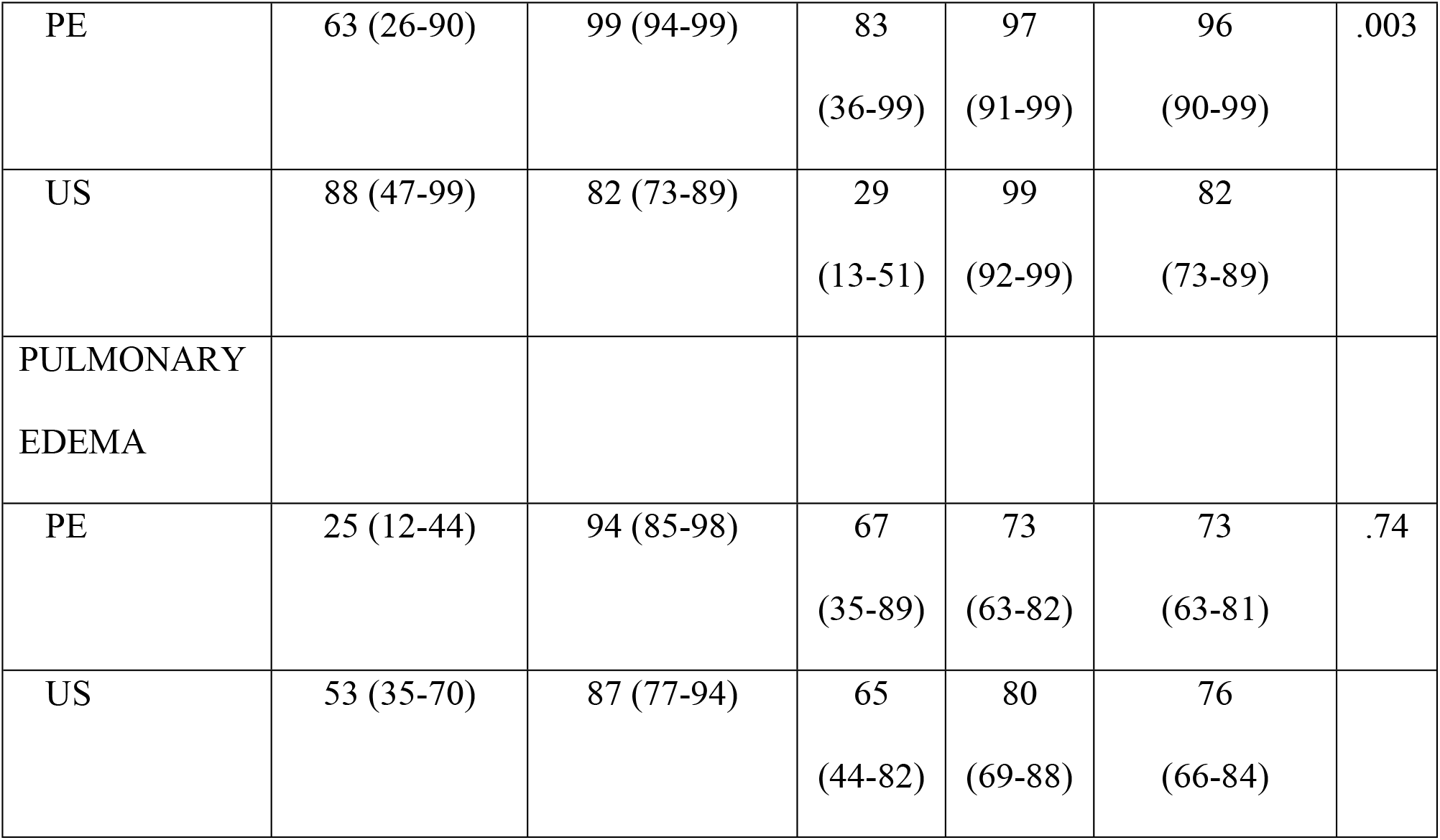
Diagnostic Characteristics by Test (Regardless of Order) CI: Confidence Intervals, PPV: positive predictive value; NPV: negative predictive value, COPD: chronic obstructive pulmonary disease, PE: physical examination, US: Ultrasound.

|  | SENSITIVITY<br>(95% CI) | SPECIFICITY<br>(95% CI) | PPV<br>(95%<br>CI) | NPV<br>(95%<br>CI) | ACCURACY<br>(95% CI) | p |
| --- | --- | --- | --- | --- | --- | --- |
| COPD |  |  |  |  |  |  |
| PE | 45 (27-64) | 86 (76-93) | 57<br>(35-76) | 80<br>(70-88) | 75<br>(65-83) | .74 |
| US | 38<br>(21-58) | 92<br>(84-96) | 65<br>(41-83) | 79<br>(69-87) | 76<br>(67-84) |  |
| ASTHMA |  |  |  |  |  |  |
| PE | 67 (24-94) | 89 (80-94) | 27<br>(9-55) | 98<br>(91-99) | 87<br>(79-93) | 1.00 |
| US | 17 (9-64) | 92 (84-96) | 11<br>(1-49) | 95<br>(88-98) | 87<br>(79-93) |  |
| PNEUMONIA |  |  |  |  |  |  |
| PE | 35 (16-59) | 80 (70-88) | 30<br>(14-53) | 84<br>(73-91) | 72<br>(62-80) | .10 |
| US | 60 (36-80) | 87 (77-93) | 52<br>(31-73) | 90<br>(81-95) | 81<br>(72-88) |  |
| PLEURAL<br>EFFUSION |  |  |  |  |  |  |
| PE | 63 (26-90) | 99 (94-99) | 83<br>(36-99) | 97<br>(91-99) | 96<br>(90-99) | .003 |
| US | 88 (47-99) | 82 (73-89) | 29<br>(13-51) | 99<br>(92-99) | 82<br>(73-89) |  |
| PULMONARY<br>EDEMA |  |  |  |  |  |  |
| PE | 25 (12-44) | 94 (85-98) | 67<br>(35-89) | 73<br>(63-82) | 73<br>(63-81) | .74 |
| US | 53 (35-70) | 87 (77-94) | 65<br>(44-82) | 80<br>(69-88) | 76<br>(66-84) |  |

***Table 4*** describes the misclassifications between COPD and pulmonary edema. There were 33 (32%) patients that had an initial diagnosis of COPD/asthma. Six of these 33 subjects in the initial L-PE group and four in the initial LUS group were later adjudicated to have a different diagnosis. Of these 10 mentioned, one in each group (two total) were later diagnosed with pulmonary edema. Of the 24 patients (24%) initially diagnosed with pulmonary edema, one patient in the initial L-PE group and seven patients in the initial LUS group were later determined to have a different diagnosis. Of these eight total cases, only one case which was in the initial LUS arm (4%) changed to COPD/asthma.

**Table 4.**
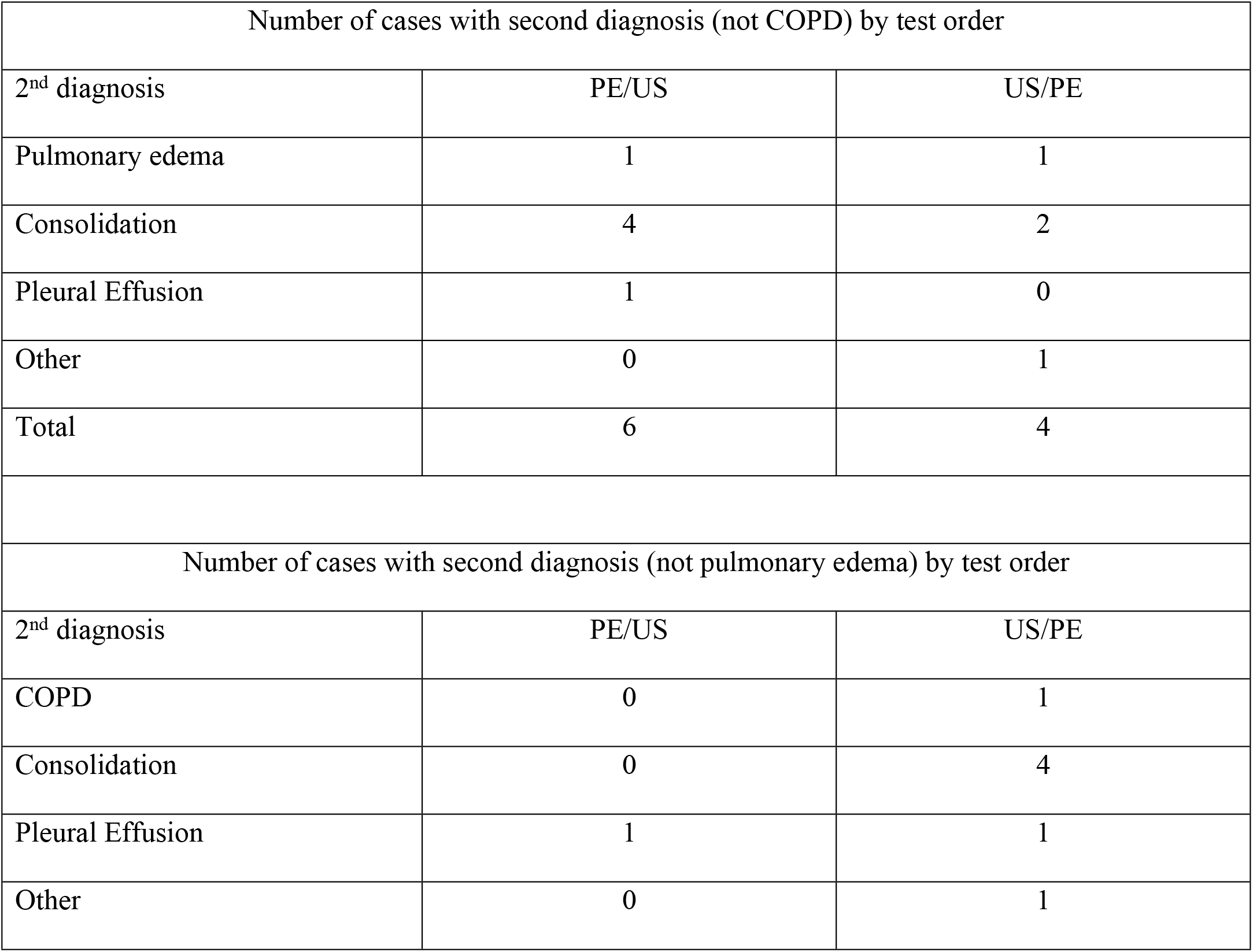
Misclassification COPD: chronic obstructive pulmonary disease, PE: physical examination, US: ultrasound.

We also stratified accuracies of LUS vs L-PE based on physician training level in ***Figure 4***. In both lower and higher training level, LUS was found to be more accurate than L-PE in diagnosing pneumonia (82% vs 73%, 78% vs 65%, respectively) and pulmonary edema (78% vs 75%, 70% vs 65%, respectively), although with no statistical significance. For COPD, lower level training physicians had higher accuracy in LUS than L-PE (77% vs 67%), but opposite was found in higher level training physicians (73% vs 78%), although with no statistical significance. For asthma, lower level training exhibited similar accuracies in either exams (84% and 84%) and there was not enough sample by higher level training physicians to provide an accuracy. For pleural effusion, L-PE was found to be more accurate than LUS in lower training level with statistical significance (95% vs 80%, 0.007) and in higher level training level although without significance (100% vs 91%).

**Figure 4.**
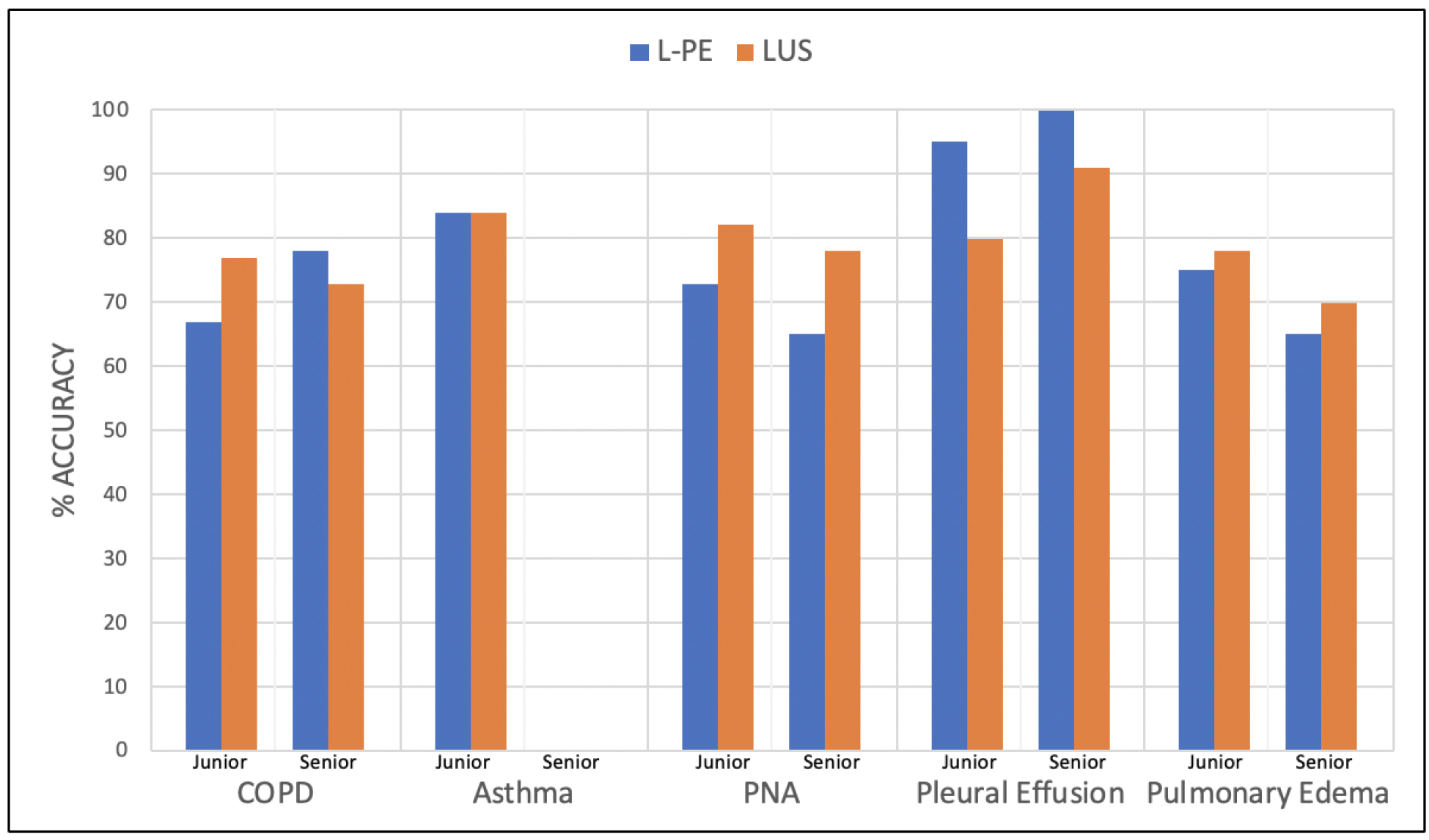
Accuracies based on physician level of training for lung physical exam (L-PE) and lung ultrasound (LUS).

### 3.3 Secondary Outcomes

Physician confidence for their leading diagnosis is noted in ***Table 5***. Physician confidence in COPD and asthma was higher for the L-PE (23% and 15% respectively) than LUS (17% and 9% respectively). Physician confidence in diagnosing pneumonia was equal between both L-PE and LUS (23% vs. 23% respectively). Physician confidence in LUS was higher than L-PE in suspecting pleural effusion (24% vs. 6%) and pulmonary edema (25% vs. 12%).

**Table 5.**
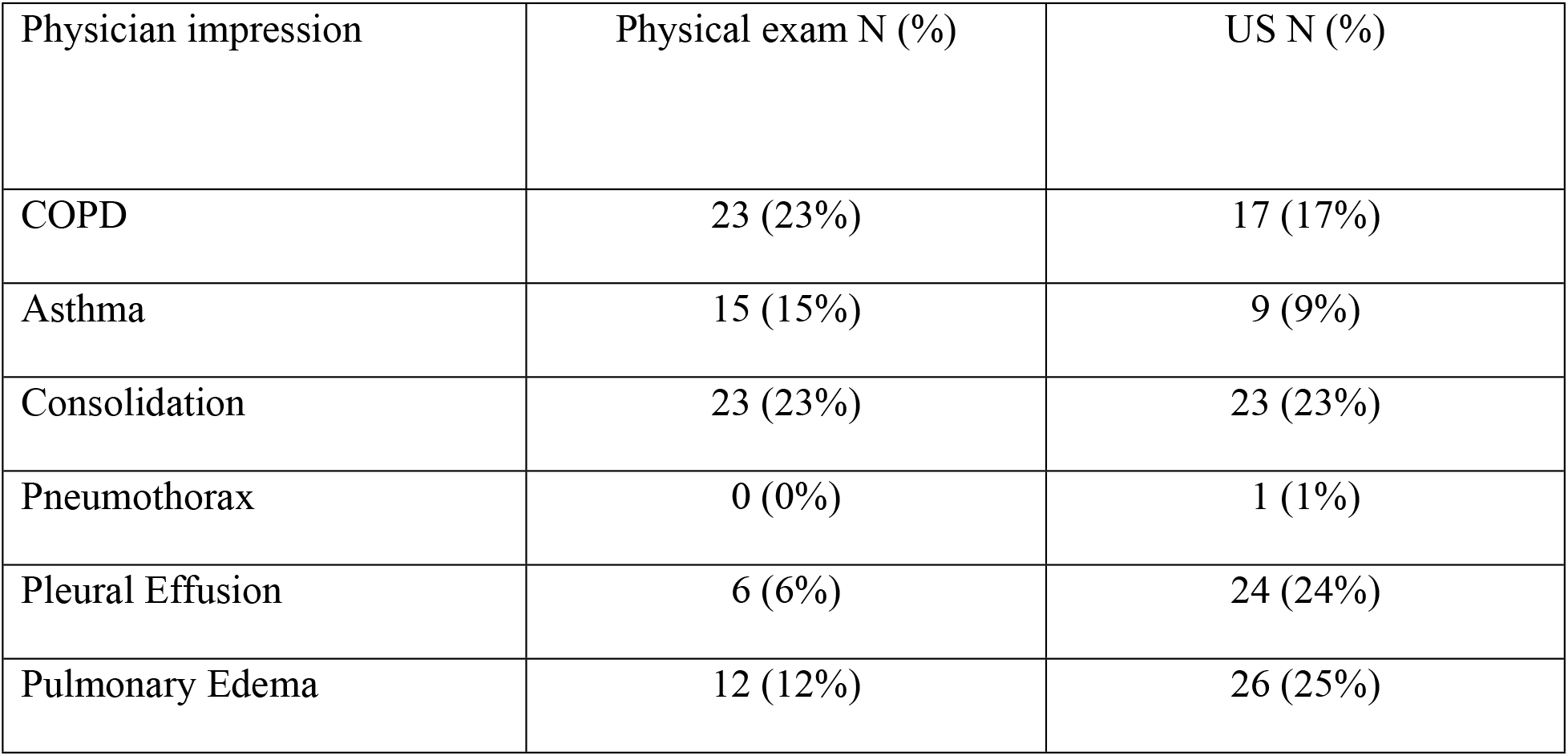
Physician impression of high confidence level by test type, regardless of order

| Physician impression | Physical exam N (%) | US N (%) |
| --- | --- | --- |
| COPD | 23 (23%) | 17 (17%) |
| Asthma | 15 (15%) | 9 (9%) |
| Consolidation | 23 (23%) | 23 (23%) |
| Pneumothorax | 0 (0%) | 1 (1%) |
| Pleural Effusion | 6 (6%) | 24 (24%) |
| Pulmonary Edema | 12 (12%) | 26 (25%) |

## DISCUSSION

This is the first study to compare lung point-of-care ultrasound to comprehensive lung physical examination in dyspneic patients presenting to the emergency department. Previous studies have compared LUS to auscultation only in respiratory conditions.^6,19,20^ The comprehensive L-PE in our study was not limited to auscultation alone, but also consisted of inspection, palpation, and percussion. As undifferentiated dyspnea consists a vast amount of etiologies, our study focused on lung pathology and lung exams and chose 6 representative respiratory conditions: asthma or COPD, consolidation, pneumothorax, pleural effusion, and pulmonary edema. Many other important causes of dyspnea such as pulmonary embolism, pericardial effusion, and ascites were not the focus of this study, although could easily be diagnosed with the incorporation of cardiac examination and bedside echocardiography. We had variable results in regard to the accuracies of comprehensive L-PE and LUS for each of six studied diagnoses. We found that LUS was slightly more accurate than L-PE for diagnosing pneumonia (82% and 72% respectively). Although not statistically significant (p=0.10), our result is consistent with prior studies in which US is a superior diagnostic tool than auscultation alone.^6,19,20^

Surprisingly, our study found L-PE to be more accurate than LUS for detecting pleural effusion (96% and 82% respectively) with statistical significance (p=0.003). This is discordant with other studies showing lung US to be more accurate than PE for this condition.^6,19,20^ However, these previous studies included only chest auscultation in their PE, whereas our study included eight comprehensive physical examination maneuvers. L-PE findings that support a diagnosis of pleural effusion include asymmetric chest expansion, dullness to percussion, and asymmetry of tactile fremitus. ^32,33^ Our inclusion of these additional PE elements might have explained this discrepancy. This suggested the possibility of a comprehensive L-PE including chest auscultation, percussion, and tactile fremitus being more accurate than LUS, but we cannot report the feasibility of performing this extensive maneuver during a busy emergency department shift. Therefore, to support the continued use of ultrasound, we cite a study by Steinmetz et al ^27^, which found the odds of correctly diagnosing pleural effusion improved by five times with addition of LUS to L-PE. Their diagnostic accuracy for pleural effusion also increased from 60 to 88% with the addition of LUS.^27^ It is therefore likely that the addition of LUS increases the diagnostic accuracy when used in combination with a thorough and comprehensive L-PE.

With regard to diagnosing COPD/asthma, our study found similar accuracies between L-PE and LUS (75% vs. 76%, p=0.74) and asthma (87%, p=1.00), with no statistical significance between the two diagnostic methods. To the best of the authors’ knowledge, there has not been any prior research directly comparing the two methods in diagnosing COPD/asthma. Previous studies have shown that the presence or absence of comet-tails (B-lines) on LUS is useful in distinguishing COPD from pulmonary edema.^22,24^ We did find LUS (76%) to be slightly more accurate than L-PE (73%) for diagnosing pulmonary edema; however, it was not statistically significant, and also our sensitivities and accuracy were found to be lower than previous literature that has shown LUS to have higher sensitivity (94%) and specificity (92%) in diagnosing cardiogenic pulmonary edema ^6,19,20,34^. This may be a result of inexperience sonographers.

As sonography is very user-dependent, stratification of data based on physician level of training was analyzed. We found that the accuracies of L-PE vs LUS in diagnosing each lung pathology did not change order. However, it is difficult to draw conclusions on physician experience when there was not enough participation by physicians with senior level of training, particularly attending participation. Also, the senior-level training group did not encounter any asthma diagnoses to allow for comparison. Therefore, it remains difficult to conclude on the effect of training level on the efficacy of these diagnostic maneuvers. Finally, we did not have large enough sample size to determine the effect of varying physician experience on the accuracy of comprehensive L-PE or LUS and should be considered in future studies.

## LIMITATIONS

There are some inherent limitations to our study. First of all, this was a convenience sample and a study performed at a single, academic institution with resident physicians performing the majority of the physical exam and ultrasound. Ultrasound is highly operator-dependent and there was not enough participation by attending physicians. To help minimize the variable, we stratified the data based on physician level of training and found similar results in this study.

Moreover, there was no blinding of the LUS group or L-PE group as this can cause potential bias in our study with the second evaluation being influenced by the first evaluation. To help minimize the amount of bias, we randomized the order of which test was performed first.

Another limitation was the considerable number (48%) of patients that had diagnoses other than the 6 pulmonary conditions we studied: bronchitis/upper respiratory infection, pericardial effusion, viral pneumonia, pulmonary embolism, chest pain/acute coronary syndrome or arrythmias. This discrepancy revealed the multitude of other medical conditions that can cause dyspnea, especially life-threatening conditions such as massive pulmonary embolism and pericardial effusion with tamponade that can be diagnosed using bedside echocardiography.^35^ These causes were considered but were not reported as we focused primarily on the lung parenchyma and did not incorporate bedside echocardiography into this particular study. In addition, a meta-analysis reports that although lung ultrasound is helpful in diagnosing pulmonary embolism, but it can detect pulmonary embolism only if the lesions extend to the periphery of lung. Therefore, it is inappropriate to use transthoracic lung ultrasonography to diagnose pulmonary embolism currently.^36^

Lastly, although previous literature demonstrates that LUS is highly accurate to diagnosis patients with pneumothorax,^37,38^ we had too small of a sample size for pneumothorax to provide the accuracy of L-PE and LUS for this condition. In addition, we did not take into consideration whether or not the patient’s body habitus affected our diagnostic abilities, nor did we ascertain the effect of patient body positioning on diagnostic accuracy of patients with B-lines for L-PE or LUS.^34^

## CONCLUSION

Neither L-PE nor LUS were statistically superior for most of the six medical conditions we evaluated, and the accuracies were overall similar. Dependent on the pulmonary condition, L-PE or LUS may be slightly more sensitive or specific over the other modality but should not be used alone. We advocate for POCUS and PE to be concurrently used, as they complement each another and maximize diagnostic accuracy of various pulmonary findings encountered in the ED. ^27^ Physicians should continue to learn and practice a good comprehensive physical exam and incorporate ultrasound to enhance their diagnostic ability.

## Data Availability

All relevant data are within the manuscript and its Supporting Information files.

## ACKNOLEDGEMENTS

The authors would like to thank Mathew Lohse, MD and Jonathan Kneib, MD, for their assistance with the study.

## REFERENCES

1. Majumdar SK. History of the stethoscope an overview. Bull Indian Inst Hist Med Hyderabad. 2002;32(2):137–150.

2. Wilkins RL. Is the stethoscope on the verge of becoming obsolete? Respir Care. 2004;49(12):1488–1489.

3. Verghese A, Charlton B, Cotter B et al. A history of physical examination texts and the conception of bedside diagnosis. Trans Am Clin Climatol Assoc. 2011;122:290–311.

4. Ünlüer EE, Karagöz A. Bedside lung ultrasound versus chest X-ray use in the emergency department. Interv Med Appl Sci. 2014;6(4):175–177.

5. Wimalasena Y, Kocierz L, Strong D et al. Lung ultrasound: a useful tool in the assessment of the dyspnoeic patient in the emergency department. Fact or fiction? Emerg Med J. 2018;35(4):258–266.

6. Lichtenstein D, Goldstein I, Mourgeon E et al. Comparative diagnostic performances of auscultation, chest radiography, and lung ultrasonography in acute respiratory distress syndrome. Anesthesiology. 2004;100(1):9–15.

7. Ferry OR, Huang YC, Masel PJ, et al. Diagnostic approach to chronic dyspnoea in adults. J Thorac Dis. 2019;11(Suppl 17):S2117–S2128.

8. Wipf JE, Lipsky BA, Hirschmann JV, et al. Diagnosing pneumonia by physical examination: relevant or relic? Arch Intern Med. 1999;159(10):1082–1087.

9. Lichtenstein DA. Lung ultrasound in the critically ill. Ann Intensive Care. 2014;4(1):1.

10. Agricola E, Arbelot C, Blaivas M, et al. Ultrasound performs better than radiographs. Thorax. 2011;66(9):828–829; author reply 829.

11. Ozkan B, Unluer EE, Akyol PY, et al. Stethoscope versus point-of-care ultrasound in the differential diagnosis of dyspnea: a randomized trial. Eur J Emerg Med. 2015;22(6):440–443.

12. Carrie C, Biais M, Lafitte S et al. Goal-directed ultrasound in emergency medicine: evaluation of a specific training program using an ultrasonic stethoscope. Eur J Emerg Med. 2015;22(6):419–425.

13. Papanagnou D, Secko M, Gullett J et al. Clinician-Performed Bedside Ultrasound in Improving Diagnostic Accuracy in Patients Presenting to the ED with Acute Dyspnea. West J Emerg Med. 2017;18(3):382–389.

14. Fakoya FA, du Plessis M, Gbenimacho IB. Ultrasound and stethoscope as tools in medical education and practice: considerations for the archives. Adv Med Educ Pract. 2016;7:381–387.

15. Geria RN, Raio CC, Tayal V. Point-of-care ultrasound: not a stethoscope-a separate clinical entity. J Ultrasound Med. 2015;34(1):172–173.

16. Fodor D, Badea R, Poanta L et al. The use of ultrasonography in learning clinical examination -a pilot study involving third year medical students. Med Ultrason. 2012;14(3):177–181.

17. Mangione S, Nieman LZ. Pulmonary auscultatory skills during training in internal medicine and family practice. Am J Respir Crit Care Med. 1999;159(4 Pt 1):1119–1124.

18. Nardone DA, Lucas LM, Palac DM. Physical examination: a revered skill under scrutiny. South Med J. 1988;81(6):770–773.

19. Inglis AJ, Nalos M, Sue KH, et al. Bedside lung ultrasound, mobile radiography and physical examination: a comparative analysis of diagnostic tools in the critically ill. Crit Care Resusc. 2016;18(2):124.

20. Vezzani A, Manca T, Brusasco C, et al. Diagnostic value of chest ultrasound after cardiac surgery: a comparison with chest X-ray and auscultation. J Cardiothorac Vasc Anesth. 2014;28(6):1527–1532.

21. Bickley LS. Bates’ guide to physical examination and history taking. Eighth edition / Lynn S. Bickley Peter G. Szilagyi. Philadelphia : Lippincott Williams & Wilkins, [2003] ©2003; 2003.

22. Lichtenstein DA, Meziere GA. Relevance of lung ultrasound in the diagnosis of acute respiratory failure: the BLUE protocol. Chest. 2008;134(1):117–125.

23. Lichtenstein DA. BLUE-protocol and FALLS-protocol: two applications of lung ultrasound in the critically ill. Chest. 2015;147(6):1659–1670.

24. Lichtenstein D, Meziere G. A lung ultrasound sign allowing bedside distinction between pulmonary edema and COPD: the comet-tail artifact. Intensive Care Med. 1998;24(12):1331–1334.

25. Lichtenstein D, Meziere G, Biderman P et al. The “lung point”: an ultrasound sign specific to pneumothorax. Intensive Care Med. 2000;26(10):1434–1440.

26. Lichtenstein D, Hulot JS, Rabiller A et al. Feasibility and safety of ultrasound-aided thoracentesis in mechanically ventilated patients. Intensive Care Med. 1999;25(9):955–958.

27. Steinmetz P, Oleskevich S, Dyachenko A et al. Accuracy of Medical Students in Detecting Pleural Effusion Using Lung Ultrasound as an Adjunct to the Physical Examination. J Ultrasound Med. 2018;37(11):2545–2552.

28. Alzahrani SA, Al-Salamah MA, Al-Madani WH et al. Systematic review and meta-analysis for the use of ultrasound versus radiology in diagnosing of pneumonia. Crit Ultrasound J. 2017;9(1):6.

29. Wang CS, FitzGerald JM, Schulzer M et al. Does this dyspneic patient in the emergency department have congestive heart failure? Jama. 2005;294(15):1944–1956.

30. Damy T, Kallvikbacka-Bennett A, Zhang J, et al. Does the physical examination still have a role in patients with suspected heart failure? Eur J Heart Fail. 2011;13(12):1340–1348.

31. Rahardjo KD, Dharmaeizar Nainggolan G et al. The diagnostic accuracy of physical examination compared to lung ultrasound for determining lung congestion in hemodialysis patients who have reached their dry weight. Journal of Physics: Conference Series. 2017;884:012150.

32. Wong CL, Holroyd-Leduc J, Straus SE. Does this patient have a pleural effusion? JAMA. 2009;301(3):309–317.

33. Diaz-Guzman E, Budev MM. Accuracy of the physical examination in evaluating pleural effusion. Cleve Clin J Med. 2008;75(4):297–303.

34. Al Deeb M, Barbic S, Featherstone R et al. Point-of-care ultrasonography for the diagnosis of acute cardiogenic pulmonary edema in patients presenting with acute dyspnea: a systematic review and meta-analysis. Acad Emerg Med. 2014;21(8):843–852.

35. Kimura BJ. Point-of-care cardiac ultrasound techniques in the physical examination: better at the bedside. Heart. 2017;103(13):987–994.

36. Jiang L, Ma Y, Zhao C, et al. Role of Transthoracic Lung Ultrasonography in the Diagnosis of Pulmonary Embolism: A Systematic Review and Meta-Analysis. PLoS One. 2015;10(6):e0129909.

37. Wilkerson RG, Stone MB. Sensitivity of bedside ultrasound and supine anteroposterior chest radiographs for the identification of pneumothorax after blunt trauma. Acad Emerg Med. 2010;17(1):11–17.

38. Staub LJ, Biscaro RRM, Kaszubowski E et al. Chest ultrasonography for the emergency diagnosis of traumatic pneumothorax and haemothorax: A systematic review and meta-analysis. Injury. 2018;49(3):457–466.

